# A national survey of potential acceptance of COVID-19 vaccines in healthcare workers in Egypt

**DOI:** 10.1101/2021.01.11.21249324

**Authors:** Aliae AR Mohamed Hussein, Islam Galal, Nahed A Makhlouf, Hoda A Makhlouf, Howaida K Abd-Elaal, Karima MS Kholief, Mahmoud M Saad, Doaa A Abdellah

## Abstract

**Background:** Since the start of COVID-19 outbreak investigators are competing to develop and exam vaccines against COVID-19. It would be valuable to protect the population especially health care employees from COVID-19 infection. The success of COVID-19 vaccination programs will rely heavily on public willingness to accept the vaccine.

**Aims:** This study aimed to describe the existing COVID-19 vaccine approval landscape among the health care providers and to identify the most probable cause of agreement or disagreement of COVID-19 vaccine.

**Methods:** A cross-sectional online survey was done.

**Results:** The present study included 496 health care employees, 55% were at age group from 18-45 years old. History of chronic diseases was recorded in 40.4%, and definite history of drug/food allergy in 10.1%. Only 13.5% totally agree to receive the vaccine, 32.4% somewhat agree and 40.9% disagreed to take the vaccine. Causes of disagreement were none safety, fear of genetic mutation and recent techniques and believe that the vaccine is not effective (57%, 20.2%, 17.7% and 16.6% respectively). The most trusted vaccine was the mRNA based vaccine. The age of health care employees and the presence of comorbidities or chronic diseases were the main factors related to COVID-19 acceptance (P<0.001 and 0.02 respectively).

**Conclusion:** Vaccine hesitancy is not uncommon in healthcare employees in Egypt and this may be an alarming barrier of vaccine acceptance in the rest of population. There is an urgent need to start campaigns to increase the awareness of the vaccine importance.

## Introduction

The COVID-19 outbreak postures a grave risk to human health (1) and investigators are competing to develop and exam vaccines against COVID-19 (2). When vaccines become available, the success of the immunization database will depend on the community’s approval of the vaccines. Recent researches have shown that although a large majority state they would agree a novel vaccine against COVID-19 (3-5), about one quarter state they would refuse (4, 6, 7).

A COVID-19 vaccine has been declared as vital to finish the pandemic and several experimental trials to evolve a vaccine for COVID-19 infection are presently being coordinated at an augmented level (1). In England, a Vaccine task-group has been developed to ‘accelerate earlier and harmonize efforts to study and then develop a corona-virus vaccine and ensure a vaccine is manufactured available to the public as early as feasible (8). The success of COVID-19 vaccination programs will rely heavily on public willingness to accept the vaccine.

Health-care employees represent an enormous number of COVID-19 contaminated individuals (9). In this circumstance, health-care employees are both probable transmitters and illness victims (10). It would be valuable to protect these health care employees from COVID-19 contagion not only for themselves, but further for their family contacts and their cases. The World Health Organization (WHO) had planned health professionals, as a significance group for COVID-19 immunization (1). The query of obligatory vaccination against COVID-19 for health professionals will be a subject of argument. After clinical development, immunization against COVID-19 will face the obstacle of community approval. WHO in 2019 identified 10 hazards for global health: including vaccine indecision and a pandemic risk (11). The world is currently fronting both hazards. Hesitancy to a vaccine refers to postponement in approval or rejection of vaccines regardless of accessibility of the services of vaccination (12). Vaccine indecision also alarms health professionals: doctors (13, 14), and nurses (15).

The aim of this study was to describe the existing COVID-19 vaccine approval landscape among the health care employees and to identify the most probable cause of agreement or disagreement of COVID-19 vaccination.

## Subjects and Methods

### Study design and sampling

A cross-sectional online-based study was conducted between 1^st^ December 2020 to 1^st^ and January 2021. The questionnaire developed on Google Forms was distributed through social media and WhatsApp groups, using the snowball technique. A total of 496 healthcare workers filled out the questionnaire that required 4 minutes to complete. Inclusion criteria were available in the consent form at the beginning of the survey. Participation in this study was voluntary, and participants received no compensation in return. The anonymity of participants was guaranteed during the data collection process.

### Minimal sample size calculation

The Epi info software (Centers for Disease Control and Prevention, Epi Info™) calculated a minimum sample of 306 participants, considering the Egyptian healthcare population 445.000 physicians, 202.542 nurses and 7000 students graduated per year (654.542 total) (16), 95% confidence level, and after adding of 4 % margin of error. A sample of 500 health care workers was targeted to allow for missing values. The final sample size included 496 participants.

### Translation and piloting

The online survey consisted of closed-ended questions in English and Arabic (for nurses, laboratory workers and radiology technicians). It was pilot tested on ten subjects to check the clarity of the questionnaire; related data were included in the final dataset. The link to Google Forms was then distributed to potential respondents.

A forward and backward translation was conducted for all the items of the questionnaire. One translator was in charge of translating the scales from English to Arabic, and a second one performed the back translation. Discrepancies between the original English version and the translated one were resolved by consensus.

### Questionnaire and data collection

An electronic questionnaire was used to collect the data. In December 2020, participants completed a questionnaire via Google forms. The aim of our sampling was to be representative of all the Egyptian health care employees based on age, sex, and profession.

### Exclusion criteria

Age below 18 years, non-health care employees and those who refuse to participate in this study.

### The following data were collected including

1. Baseline demographic data including the occupation status and the presence of comorbid disorders.
2. Query whether the participant had history of allergy to certain food or drugs.
3. Question whether the participants, their families or friends had previous attack of COVID-19.
4. Question about the intention of health care employees to take COVID-19 vaccine in a 5-point Likert Scale: (1 = totally disagree; 5 = totally agree).
5. Query about the desirable type of COVID-19 vaccine whether Pfizer, Russian, Chinese, Moderna or Oxford/AstraZeneca subtypes.
6. Participants were asked about their source of information about COVID-19 vaccine comprising social media sources and the TV.
7. Question about the cause of refusal to take COVID-19 vaccine either not clinically safe, not effective, theory of genetic mutation, fear of recent Nano-technology, no indication for the vaccine, or the concept that the subject already had a previous attack of COVID-19 and had immunity.
8. Lastly, the participants were asked if they had received the seasonal influenza vaccine in a 5-point Likert Scale: (1 = never; 5 = regularly taken every year).
9. Participants provided informed consent prior to data collection.

***The study was approved*** by the ethical committee Assiut Faculty of Medicine and registered in ClinicalTrials.gov Identifier: NCT04694651

### Statistical analysis

Completed forms were imported into a Microsoft Excel spreadsheet. Data were then analyzed on Statistical Package for the Social Sciences (SPSS) software version 23 (Chicago, IL, USA). A descriptive analysis was performed using the counts and percentages for categorical variables and means and standard deviations for continuous measures. bivariate analysis was used to assess the factors that can affect the acceptance of the vaccine. The statistical significance was set at a p-value <0.05.

## Results

The present study included 496 health care employees, 34.9% male and 65.1% female, 55% were at age group from 18-45 years old and only 0.4% more than 65 years old. Only 17.8% had history of COVID-19 infection but 62.1% reported COVID-19 in their family/friends. History of chronic diseases was recorded in 40.4%, and definite history of drug/food allergy in 10.1% **(*Table 1*)**.

**Table 1.**
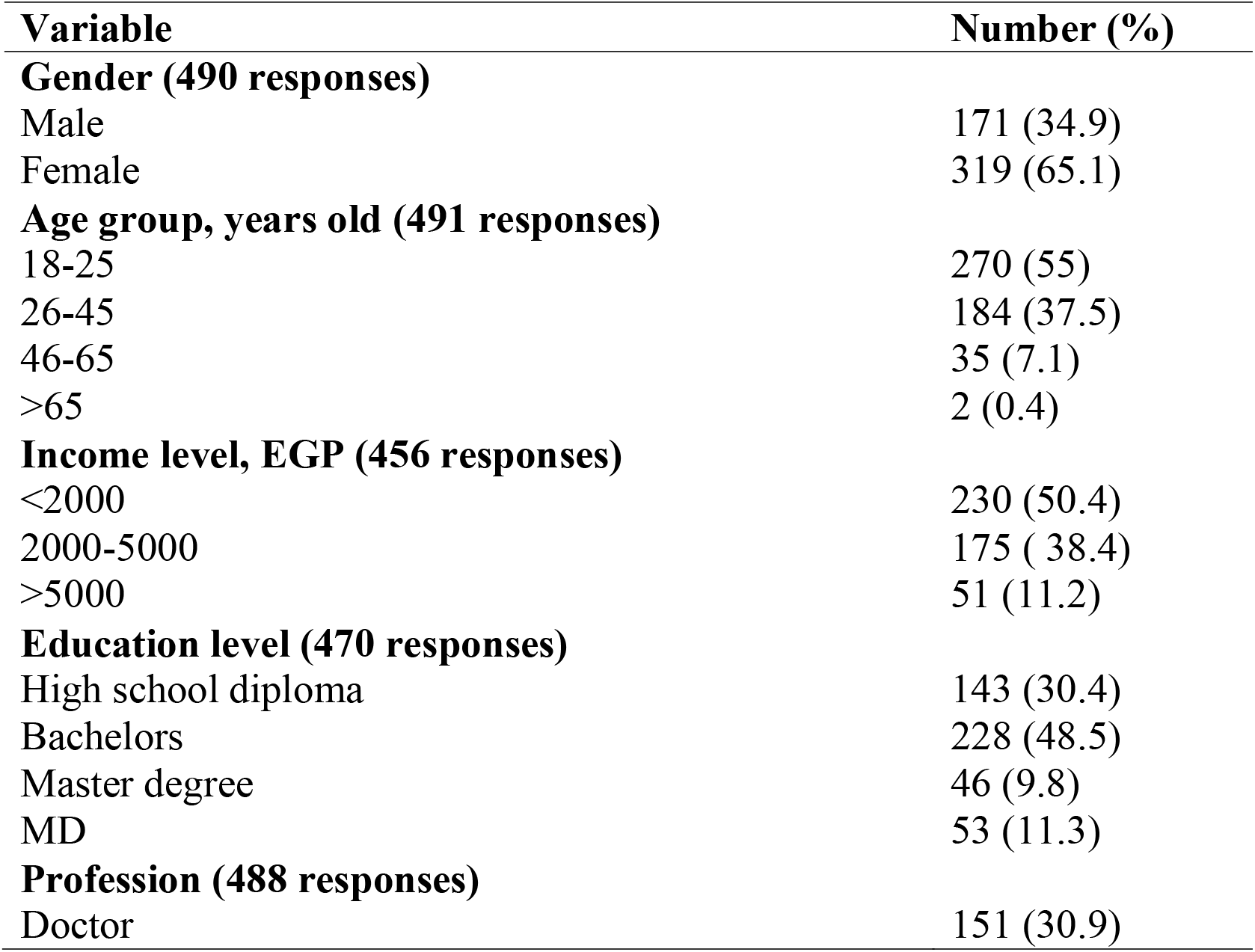
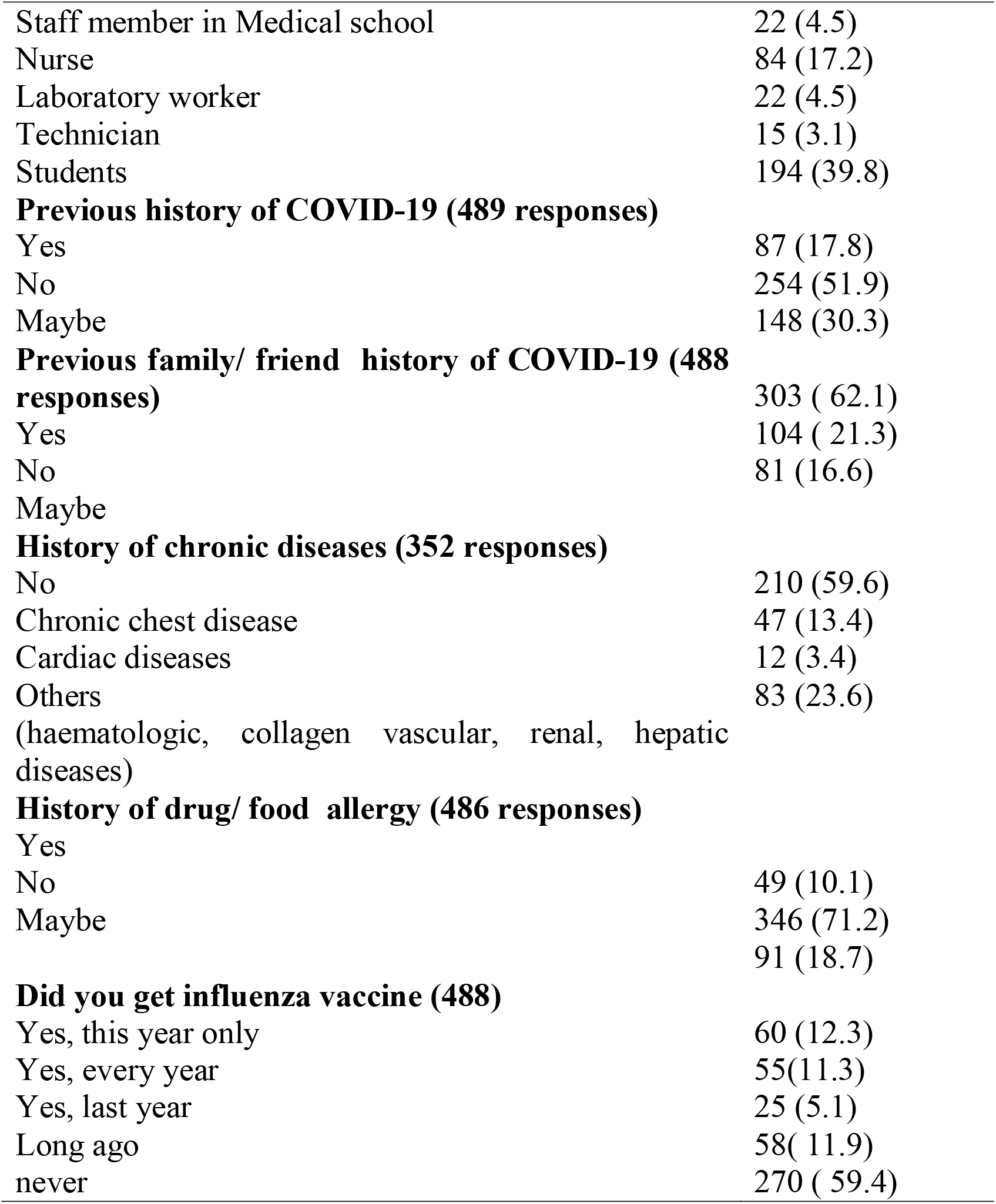
Descriptive data of health care employees included in the survey (n=496)

***Table 2*** showed that only 13.5% totally agree, 32.4% somewhat agree and 13.2% had no opinion. However, 40.9% disagreed to take the vaccine. Causes of disagreement were not safe, fear of genetic mutation and recent techniques and believe that the vaccine is not effective (57%, 20.2%, 17.7% and 16.6% respectively). The most trusted vaccine was the mRNA vaccine developed by Pfizer/BioNtec.

**Table 2.**
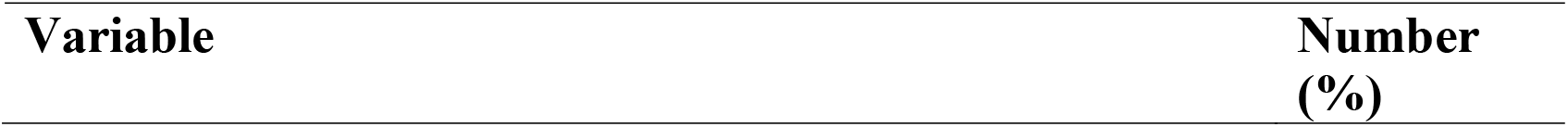
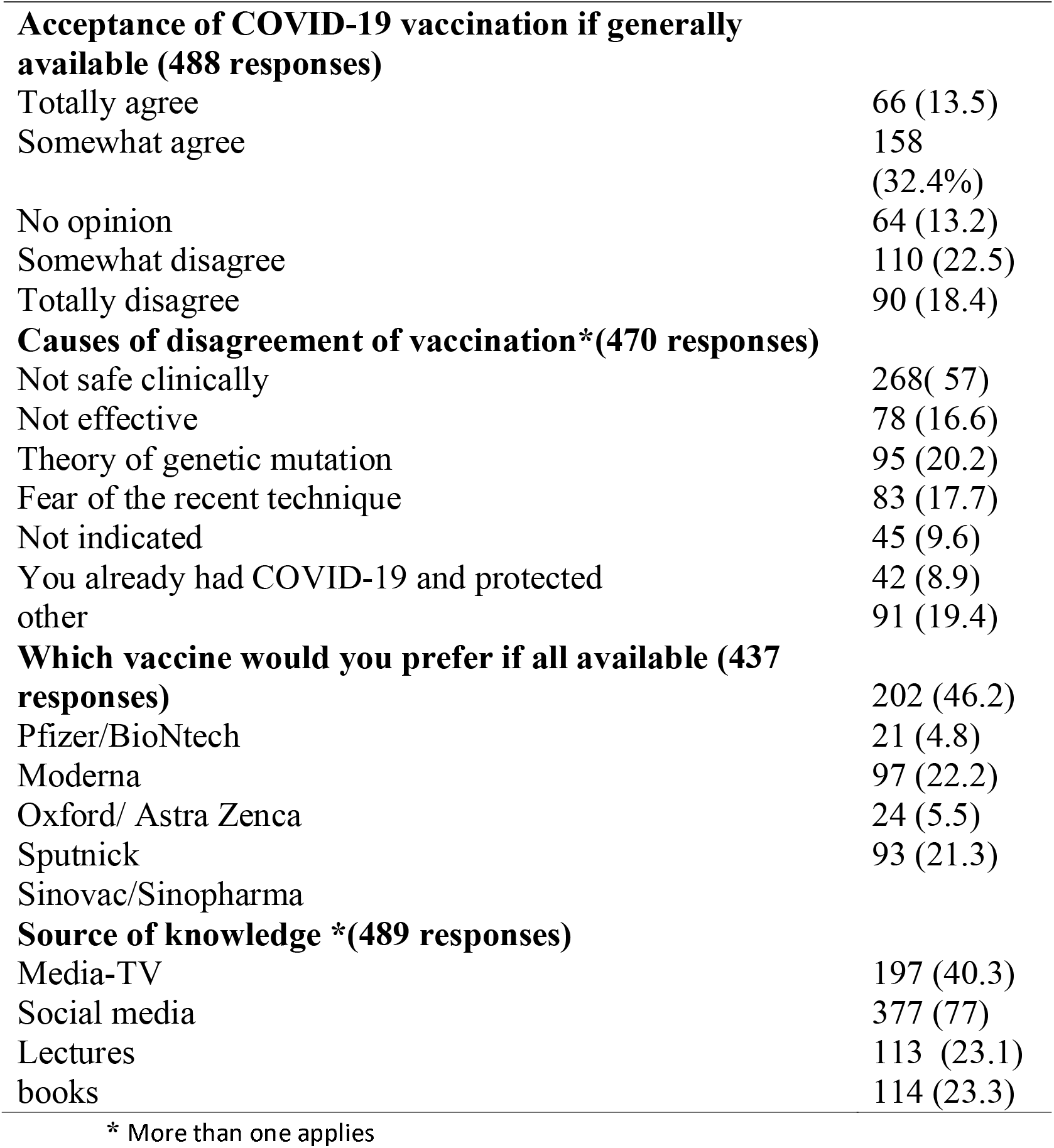
Acceptance and attitudes about the COVID-19 vaccination.

To examine the factors determining the acceptance or refusal of vaccine, a Bivariate analysis showed that the age of health care employees and the presence of comorbidities or history of chronic diseases were the main factors related to COVID-19 acceptance (P<0.001 and 0,02 respectively) **(*Table 3*)**.

**Table 3.** Bivariate analysis of vaccine acceptance in different groups (n=409)

| Variable | Value | Asymp. Sig (2-sided) |
| --- | --- | --- |
| **Gender** |  |  |
| Pearson Chi-Square | 5.457<sup>a</sup> | 0.065 |
| Likelihood Ratio | 5.466 | 0.065 |
| **Age** |  |  |
| Pearson Chi-Square | 287.198 | **0.001\*** |
| Likelihood ratio | 64.960 | **0.001\*** |
| **Education level** |  |  |
| Pearson Chi-Square | 4.386 | 0.62 |
| Likelihood Ratio | 4.797 | 0.57 |
| **Income** |  |  |
| Pearson Chi-Square | 1.394 | 0.845 |
| Likelihood Ratio | 1.773 | 0.777 |
| **Profession** |  |  |
| Pearson Chi-Square | 10.072 | 0.434 |
| Likelihood Ratio | 10.852 | 0.369 |
| **Previous COVID-19 infection** |  |  |
| Pearson Chi-Square | 3.810 | 0.432 |
| Likelihood Ratio | 3.171 | 0.530 |
| **History of Chronic diseases** |  |  |
| Pearson Chi-Square | 27.797 | 0.06 |
| Likelihood ratio | 31.783 | **0.02\*** |
| **History of previous drug/food allergy** |  |  |
| Pearson Chi-Square | 5.453 | 0.244 |
| Likelihood Ratio | 4.673 | 0.323 |

## Discussion

The current study surveyed the potential acceptance of health care employees to COVID-19 vaccines as they serve on first line of pandemic response efforts and more exposed to infection. The main findings showed that 45.9% agree to receive the vaccine (13.5% totally agree, 32.4% somewhat agree) and 40.9% disagreed to take it. Causes of disagreement were none safety, fear of genetic mutation and recent techniques and believe that the vaccine is not effective. The age of health care employees (older participants tend to approve more) and the presence of comorbidities or chronic diseases were the main factors related to COVID-19 acceptance.

During 2020, the international organizations and national regulatory authorities in collaboration with multinational pharmaceutical industry directed their efforts for developing COVID-19 vaccines as the pandemic continues. By the end of 2020 five vaccines have been authorized to be used on emergency use authorization (EUA) basis (17). In the current study, 32.4% of participants expressed their response using statement “somewhat agree” to accept the vaccine while the lower percentage of participants 13.5 % used “totally agree”. These indicate general willingness of health care employees to accept the vaccine, however “somewhat agree” statement may indicate hesitancy and background specific concern regarding new vaccine acceptance. This hesitancy was confirmed when the participants were asked about the causes of disagreement of vaccination. The higher percentage 57% of participants said “not safe clinically” which indicate lake of reassurance regarding vaccine safety and future unknown adverse events.

In the context of the vaccine acceptance, a survey study was done in Congo included 613 Congolese health care workers (HCWs) reported only 28% of participants would accept the vaccine against COVID-19 (18). Another study from France included 3259 respondents to the online questionnaire and it was observed that nearly 3/4 of the participants (77.6%, 95% CI 76.2–79%) would accept the vaccine. The proportion of healthcare workers willing to get vaccinated was 81.5%, while it was 73.7% in non-healthcare workers (4). Moreover, studies conducted in USA regarding intention to accept the vaccine reported that only 30% of participants would not like to receive the vaccine soon when it becomes available (19, 20).

The intention to accept the vaccination against COVID-19 among the Egyptian HCWs is low compared to studies from western countries but better than African’s; this may be interpreted by many factors: misinformation that gained from social media as a source of knowledge for the participants (21). Second; the variation of the profession of the respondent in this study may be another factor reflected on our results as high percentage of respondents were students at medical school (39.8%) who have less level of knowledge that surly different from doctors. Third: the time factor can be added to the difference between results of the present study and the other compared studies as most of these studies were conducted early in 2020 (March to May). It was the peak of pandemic globally and it would believe that the vaccine is the magical solution to control infection that can affect the decision of the respondents participated in the different studies.

Regarding the type of the vaccine it was demonstrated that 46.2% of participants prefer mRNA based vaccine (Pfizer/BioNtech) vaccine if available while the remaining 53.8% were distributed between the other types of the vaccine. It can’t sure why Pfizer vaccine more preferable to be accepted by the participants more than other types of vaccine, but it may relate to the trust to this brand and transparency of information presentation regarding their vaccine in public.

The current results demonstrated that social media and TV media are the main source of knowledge for the participants; 77 % and 40.3%, respectively. Unfortunately these sources are not preferable to be the source of knowledge due to misinformation that can they give to the public (conspiracy theory) such as some social media posts claimed the use of mRNA based vaccine for COVID-19 can change population’s DNA (21, 22). This is may be added to increase the hesitancy of vaccine acceptance.

To assess the factors that can affect the acceptance of the vaccine in present study, univariate regression analysis was used, and the study demonstrated significant correlation regarding age and history of chronic diseases. This may be explained by fear of those groups about the impact COVID-19 on their comorbidities reported in many studies indicating that Diabetes Mellitus (23) chronic hepatic and renal diseases (24) and multiple comorbidities especially neurologic can increase both morbidity and mortality in COVID-19 patients (25).

According to the present study, although the higher percentage of the participants had the intention to accept the vaccine but overall, it considers low acceptance of the respondents. The vaccine hesitancy in the Egyptian HCWs can be the major barrier that affects the decision of the vaccine acceptance in Egypt. According to a global survey conducted in

19 countries included 13,426 participants to assess the potential acceptance of COVID-19 vaccine; 71.5% would somewhat like to uptake the vaccine. Although they reported the higher percentage of vaccine acceptance but differences in the acceptance ranged from (80%) in Asian nations and less than (55%) in Russia (26). Accordingly, it is not surprising that acceptance of the vaccine is low in Egypt as vaccine hesitancy exist globally.

The concerns about the safety of the vaccine was recorded in 57% of participants in the study. Similar data were reported in different areas and numerous factors associated with vaccine hesitancy all over the world included a lot of inquiries about safety and effectiveness of the vaccine (27), which lasted even since the SARS-CoV-1 outbreak (28). Fear from rapid release of the vaccine to public with lack of adequate research, lack of research on Arab population lead increase uncertainty about the value of receiving the newly developed vaccines.

## Conclusion

Vaccine hesitancy is not uncommon in healthcare employees in Egypt and this may be an alarming barrier of vaccine acceptance in the rest of population. There is an urgent need to start campaigns to increase the awareness of the vaccine importance.

## Data Availability

Data available on request

## Recommendations

There is a need to correct the false concepts and misinformation that taken from social media background. Building confidence track by clear communication between national government officials and HCWs. All of this can be achieved by explanation of how the vaccine is work, level of effectiveness, safety and expected adverse events and method of the vaccine uptake (doses and site). Lectures preparation should be given by trusted leaders in medical field to HCWs and to answer all inquiries in their mind.

## Acknowledgement

none

